# Genomic Profiling to Contextualize the Results of Intervention for High-Risk Smoldering Myeloma

**DOI:** 10.1101/2023.08.30.23294483

**Authors:** Dickran Kazandjian, Benjamin Diamond, Marios Papadimitriou, Elizabeth Hill, Romanos Sklavenitis-Pistofidis, Bachisio Ziccheddu, Patrick Blaney, Monika Chojnacka, Michael Durante, Kylee Maclachlan, Ryan Young, Saad Usmani, Faith Davies, Gad Getz, Irene Ghobrial, Neha Korde, Gareth Morgan, Francesco Maura, Ola Landgren

## Abstract

Early intervention for High-Risk Smoldering Multiple Myeloma (HR-SMM) achieves deeper and more prolonged responses compared to Newly Diagnosed (ND) MM. It is unclear if beneficial outcomes of interventional studies in HR-SMM are due to treatment of less complex, susceptible disease or inaccuracy in clinical definition of cases entered. Here, to gain greater biologic insight into treatment outcomes, we performed the first whole genome sequencing analysis of treated HR-SMM for 27 patients treated with carfilzomib, lenalidomide, and dexamethasone and lenalidomide maintenance (NCT01572480). Genomic features were pooled with another contemporary HR-SMM interventional study (E-PRISM; NCT02279394) and compared to those of NDMM. We reveal that across interventional cohorts, the genomic landscape of HR-SMM is uniformly simple as compared to NDMM counterparts, with fewer inactivation events of tumor suppressor genes, fewer RAS pathway mutations, lower frequency of *MYC* disruption, and lower APOBEC contribution. The absence of these genomic events parallels that of indolent precursor conditions with low chance of progression, possibly explaining the overall superior outcomes across these trials. However, there remains a subgroup of patients harboring genomic complexity for whom early intervention with potent triplet therapy fails to sustain response and who experience resistant, progressive disease. Overall, these results suggest that clinical risk scores do not effectively discriminate between genomically indolent and aggressive disease. Furthermore, our study supports the use of genomics to contextualize the advantage of early intervention in SMM and to consider novel approaches for those with the most aggressive precursor states.

**Key Points:** Treated clinical high-risk smoldering multiple myeloma is genomically heterogeneous but is mostly less complex than multiple myeloma counterparts.

A small subgroup of high-risk genomic features is associated with disease progression despite early intervention with triplet therapy.

## INTRODUCTION

Multiple Myeloma (MM) is a plasma cell neoplasm that progresses through precursor states before evolution to symptomatic cancer^1-6^. On the spectrum of precursor conditions is smoldering MM (SMM); a clinically-defined state of asymptomatic expansion of clonal plasma cells for which approximately 60% of affected individuals will experience progression to overt disease within 10 years of diagnosis^1, 3^. Various risk models have been developed to stratify the risk of progression and are thus far based on surrogate clinical markers and indirect measures of disease burden^6-9^. It has been hypothesized that by intervening before evolution to clinically significant disease (i.e., MM), therapeutic outcomes might be improved by preventing organ damage and by treatment of a less expanded, less complex disease state. Based on these principles, multiple investigations have been developed for treatment of individuals with SMM at high risk (HR-SMM) of progression to MM ^6, 10-13^. In the era of modern drugs, early intervention for HR-SMM has consistently resulted in deeper and more prolonged treatment responses compared to newly diagnosed MM (NDMM), often with less intense therapy. However, it is unknown if this benefit is due to treatment of a less complex, susceptible entity in a fit individual or inaccuracy and incongruity with and between differing risk models resulting in treatment of more biologically indolent disease^14^.

The introduction of next generation sequencing (NGS) technologies has promoted a gradual shift from the use of clinical surrogate markers of disease burden to genomic determinants of disease biology^5, 15-17^. Whole genome sequencing (WGS) and whole exome sequencing (WES) have expanded upon and delineated several myeloma-defining genomic events^18-22^. These events include presence of complex structural variants (SV), APOBEC mutational signatures, mutations involving the MAPK and NFkB pathways, loss of tumor suppressor genes, and events involving MYC^17^. However, the genomic landscape of clinical HR-SMM that has been subjected to early therapeutic intervention has not been comprehensively explored.

To contextualize the superior outcomes of contemporary SMM treatment trials, we performed the first WGS-based analysis of patients with HR-SMM, using samples and data from a cohort of HR-SMM patients treated with carfilzomib, lenalidomide, and dexamethasone and lenalidomide maintenance (KRd/R) as part of a phase II clinical trial with long-term clinical follow-up^10^. Furthermore, in our analysis we included information from an additional HR-SMM phase II interventional trial (E-PRISM) and we evaluated genomic lesions and clinical outcomes across the two studies, and compared against those of NDMM. Overall, we confirm wide discordance among different SMM prognostic models for MM progression, but a common genomic simplicity that provides the biologic rationale for the favorable responses observed on interventional trials. Conversely, patients harboring complex genomic profiles and distinct known MM genomic drivers generally failed to achieve sustained negativity for minimal residual disease (MRD) and experience progression of disease, suggesting that early triplet-therapy intervention does not completely overcome the adverse impact of these features on disease evolution and clinical outcomes.

## METHODS

To gain biologic insight into treatment outcomes, we performed WGS of baseline samples for 27 treated patients with HR-SMM. Patients received 8 cycles (32 weeks) of carfilzomib 20/36 mg/m^2^ on days 1, 2, 8, 9, and 15; lenalidomide 25mg days 1-21; and dexamethasone 20mg twice weekly for cycles 1-4 and 10mg for cycles 5-8; followed by 2 years of maintenance lenalidomide (KRd/R; NCT01572480)^10^. Clinical records were updated from prior publication and reviewed to correlate trial outcomes with genomic features. We pooled genomic features with whole exome sequencing data (WXS) from a second cohort of 27 patients with HR-SMM treated with Elotuzumab-R+/-d (Elo-Rd; E-PRISM; NCT02279394)^13^ and compared to 701 patients with NDMM from CoMMpass (NCT01454297) with available WGS and WXS. Additional comparators included WGS from 60 patients with NDMM treated with daratumumab(dara)-KRd (NCT03290950) and KRd (NCT02937571)^23, 24^. Samples and data were obtained and managed in accordance with the Declaration of Helsinki. Sample IDs were uniquely generated and were not known to anyone outside the research group

All patients had protocolized testing for MRD yearly. We considered any patient achieving MRD-negativity on trial as having MRD-negativity as a best response. We considered sustained MRD-negativity as at least two consecutive MRD-negative measurements at least 12 months apart. For the purposes of our analysis, for a patient to be considered to have sustained MRD-negativity, their MRD measurement at the most recent follow-up visit must have remained negative. Finally, we considered both biochemical progression and progression to overt MM requiring treatment as progression of disease.

Each specific statistical test is annotated in the text. Fisher’s Exact Test and the Wilcoxon Ranked Sum test were used to compare differences between groups. P-values <0.05 were considered statistically significant. Survival data were analyzed and visualized with Kaplan-Meier methods.

A detailed description of the sequencing and analytical methods is provided in the **Supplemental Methods**.

## RESULTS

### Clinical outcomes for patients with HR-SMM treated with KRd/R

Clinical features for the KRd/R cohort are listed in **Supplemental Table 1**. After a median follow-up of 52.8 months, median PFS was unreached for patients treated with KRd/R (**Figure 1a-b**). After 8 cycles of KRd, 19 (70.3%) achieved MRD-negativity (Flow-MRD; LOD 10^−5^, **Figure 1a**). For comparative reference, acknowledging the pitfalls of comparison across trials, patients with NDMM treated with KRd achieved end-induction MRD rates of 56% (FORTE; 48 weeks KRd).^25^ At time of data cutoff, June 13, 2023, 14 patients (51.9%) achieved sustained MRD-negativity at last follow up, 6 patients (22.2%) lost an initial MRD-negative response and 5 patients experienced progression of disease [18.5%; 4 biochemical and 1 progression to MM (NIH018A; **Supplemental Table 1, Methods**)].

**Figure 1:**
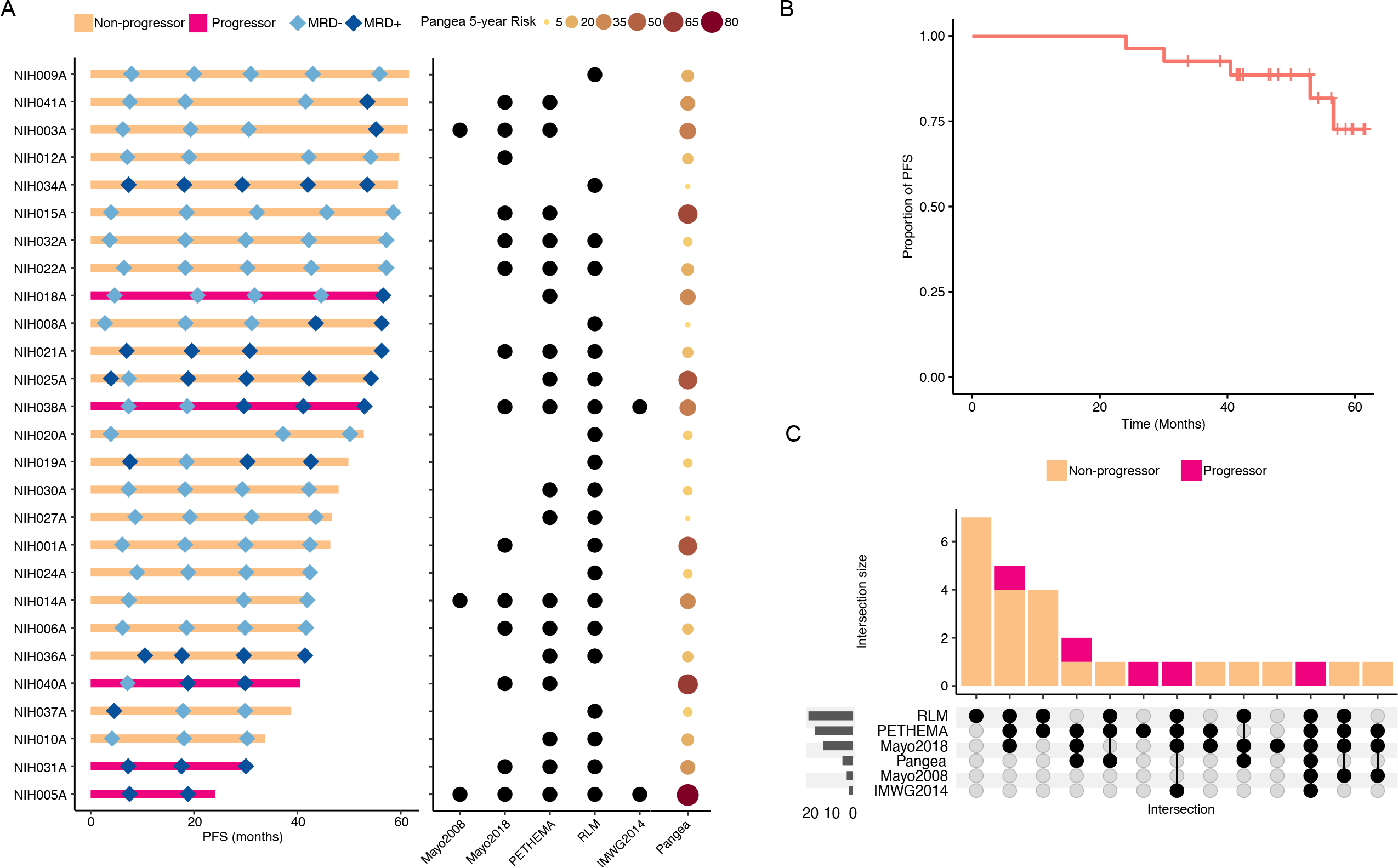
Clinical summary of the KRd/R cohort. **A**. Swimmer plot of progression-free survival (PFS) annotated with serial MRD measurements (left panel) and high-risk status by relevant clinical risk scores (right panel). **B**. PFS curve for the KRD/R cohort generated with Kaplan-Meier methods. **C**. Upset plot demonstrating incongruity between clinical risk scores as they pertain to outcomes after treatment with KRD/R.

Accrual on NCT01572480 began in 2012 and, as such, 2 patients would today be reclassified as MM per 2014 IMWG criteria on the basis of bone marrow plasma cells ≥60%^26^. Both of these patients (NIH005A and NIH038A) experienced disease progression on study. Otherwise, 3 (11.1%) were HR by Mayo2008 criteria, 14 (51.9%) retrospectively by Mayo 20/2/20, 18 (66.7%) by PETHEMA, and 21 (77.8%) by Rajkumar/Landgren/Mateos criteria^6-9^. 18 (66.7%) met criteria by 2 or more scores (**Figure 1c**) and the median 5-year risk of progression per the Pangea model was 18.6% (range 4.8-82.1)^27^. Overall, though all patients on trial were considered high-risk, there is clear discordance between competing clinical risk models and a wide range in predicted risk of progression^14^.

### The genomic landscape of treated HR-SMM

To contextualize the overall favorable outcomes of treated HR-SMM, we sought to qualify genomic features across studies by pooling WGS data with available WXS from the E-PRISM study^13^. Genomic data from treated HR-SMM were then compared to 701 patients with NDMM from the CoMMpass study. As an initial check for bias towards less aggressive disease, there were no differences in the frequency of high-risk FISH-based cytogenetic lesions [(gain1q, amp1q, t(4;14), t(14;16), del(17p)] between treated HR-SMM and NDMM.

Within the KRd/R HR-SMM WGS cohort, the median mutational burden was 4134 (range, 2021-8346), which was significantly lower compared to WGS from NDMM treated with KRd +/- dara (median, 5179; range, 1157-14471, p = 0.019). To investigate differences in mutational processes involved in HR-SMM and NDMM we profiled the mutational signatures landscape of the KRd/R cohort (**Figure 2a, Supplemental Table 2**), NDMM treated with KRd +/- dara (**Supplemental Figure 1**), and WXS from the E-PRISM cohort (**Figure 2b**). Overall HR-SMM contained the same mutational signatures detected in NDMM and in prior studies of myeloma precursor disease (**Supplemental Methods**) ^17, 21^. However, HR-SMM patients enrolled in both the KRd/R and the E-PRISM studies had a significantly higher proportion of patients without evidence of APOBEC (SBS2+SBS13) mutagenesis compared to NDMM [WGS 48.1% vs 86.7%, p < 0.001; WXS 14.8% vs 45.2% (CoMMpass), p < 0.001; **Figure 2C-D**]. Similarly, cases of hyper-APOBEC mutagenesis (SBS2+SBS13>11%; **Supplemental Methods**), which has been associated with poor prognosis in MM,^28^ were under-represented in HR-SMM (p = 0.035). Overall, these findings are consistent with prior observations that APOBEC activity is lower in SMM and increases across the continuum of progression to MM^17, 28^. Importantly, however, APOBEC activity has been identified in over 82-85% of myeloma precursors destined for progression and its relative absence here is suggestive that despite clinical high-risk status, many of these cases may represent more indolent conditions^17^.

**Figure 2:**
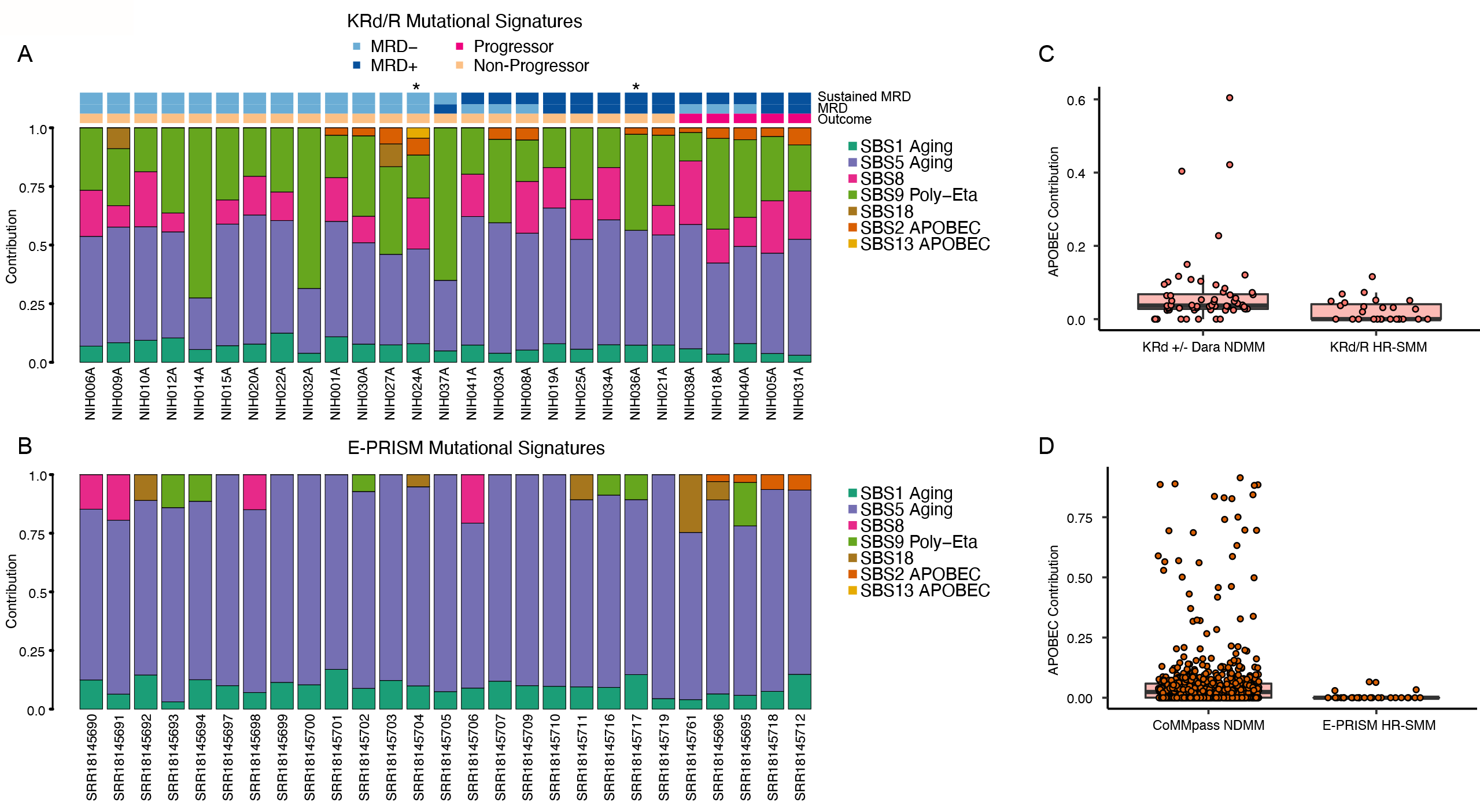
Mutational Signatures Landscape of treated HR-SMM. **A**. Mutational signatures contribution for each case treated with KRD/R. Asterisks denote MAF(B)-translocated cases. SBS: Single Base Substitution. **B**. Mutational signatures contribution for each case treated on E-PRISM. **C**. Comparison of APOBEC (SBS2 + SBS13) contribution between NDMM treated with (dara)-KRd vs. HR-SMM treated with KRd/R. **D**. Comparison of APOBEC contribution between NDMM from CoMMpass (i.e., WXS calls) vs. HR-SMM treated on E-PRISM.

We next interrogated the landscape of driver events across the cohorts using a catalog of 91 known genes involved by mutations and focal copy number aberrations derived from de novo driver discovery across 1933 NDMM^29^. We then combined mutations in driver genes with copy number aberrations (CNA; **Supplemental Figure 2**), including known GISTIC hotspots^20, 29^, to define inactivation of tumor suppressor genes (**Supplemental Figure 3, Supplemental Methods**). Consistent with their later role in MM evolution^16, 30^, a large number of tumor suppressor genes were less frequently inactivated, including *CDKN2C, CYLD, TENT5C, FUBP1, MAX, NCOR1, NF1, NFKBIA, PRDM1, RB1, RPL5*, and *TRAF3* (p < 0.05; **Supplemental Table 3**). Restricting scope only to biallelic inactivations of tumor suppressor genes, *TRAF3* was more frequently inactivated in MM (69/701 vs 1/54; p = 0.05) and importantly, the only HR-SMM case (NIH038A) would be reclassified as MM with current IMWG criteria. This finding reinforces the role of the NF-ΚB pathway as a late driver in the pathogenesis of MM^31^. Consistent with their later role in the genomic evolution from SMM to MM, mutations in RAS pathway genes (*NRAS, KRAS, BRAF*; p < 0.001) and amplifications at 8q24 (i.e., *MYC*; p = 0.03) were more frequent in MM^17, 32^. Finally, sensitivity analysis was performed by removing both cases that would today be reclassified as MM per IMWG 2014 criteria^26^, with little change in observations (**Supplemental Tables 3 and 4**).

For the available full coverage WGS samples (KRd/R HR-SMM and KRd +/- dara NDMM) we compared the landscape of structural variants (SV). There were no significant differences in the frequency of involvement of SV hotspots between HR-SMM and NDMM. Though there was not a significant difference in the frequency of chromothripsis events in the KRD/R genomes, alone, using CN signatures to estimate the presence of chromothripsis from the E-PRISM WXS, there was a lower frequency of chromothripsis events in pooled HR-SMM (p = 0.048), consistent with the SV’s association with complex genomes and poor outcomes **(Supplemental Methods, Supplemental Figure 4 A-B**)^20, 33^.

Overall, we see that across cohorts of HR-SMM, genomic features associated with NDMM (i.e., Myeloma-defining genomic events) are significantly underrepresented. As clinical HR-SMM should theoretically be at risk for imminent progression to MM (i.e., 2-5 years), most genomic drivers should already have been acquired on its course to overt disease^17, 19, 32^. This suggests that clinical risk scores, based on surrogate markers of disease burden, have limitations in accurately recapitulating the underlying disease biology that contributes to disease aggression and imminent progression and that their use in trial settings may lead to inadvertent capture of indolent disease states.

### Genomic features associated with outcomes HR-SMM treated with triplet therapy

We next sought to investigate the genomic lesions associated with clinical outcomes following intervention with potent triplet therapy. Sustained MRD-negativity and disease progression (either biochemical or clinical) during intervention with KRd/R were the key endpoints. Deletions of chromosome 13q14.2 (*RB1*) and inactivation of *MAX* and *HIST1H2BK* (p < 0.05) were each associated with disease progression, as was any presence of APOBEC mutagenesis (p = 0.016; **Supplemental Table 5**). Examining SV, chromothripsis, known to be associated with poor outcomes in MM, was likewise seen here to be a risk factor for disease progression (p = 0.030) as were SV involving *TENT5C* (p=0.028; **Supplemental Methods, Supplemental Table 4**)^20^. In fact, progressors were more likely to have a higher number of involved SV hotspots, signifying the role of co-occurrence of multiple disease drivers (i.e., genomic complexity) as a main contributor to therapy resistance and continued clinical disease evolution (**Supplemental Figures 4A-C**).

We next examined genomic features within the context of clearance of MRD following treatment with KRd/R. No genomic features emerged as significantly associated with achievement of MRD-negativity following 8 cycles of KRd (primary endpoint). On the KRd/R study, patients had protocolized yearly MRD testing and we observed that gain of 1q, inactivation of *MAX*, deletion of 14q24.3, and t(4:14) were each associated with failure to sustain MRD-negativity at last follow-up (**Supplemental Table 4**).

As HR-SMM is a clinical definition and given the wide heterogeneity and relatively simplicity of the genomic landscape of most of the HR-SMM enrolled in KRd/R and E-PRISM, we sought to determine what genomic lesions shared with NDMM were more closely representative of biologically progressive, clonally mature disease. In an orthogonal approach, we removed clinical annotations and performed hierarchical clustering of genomic features from all 701 NDMM from CoMMpass and all 54 HR-SMM (**Supplemental Figure 5, Supplemental Methods**). In parallel to the prior clinical comparisons, APOBEC activity, gain1q, and loss of tumor suppressor genes all clustered together and often co-occurred as features of genomic complexity that predict poor outcomes in terms of PFS and sustained MRD-negativity. Furthermore, the majority of cases enrolled in the KRd/R HR-SMM clustered within the most indolent and genomically uncomplicated group of NDMM.

Finally, a summary of all significant genomic features was compiled and considered (**Figure 3A**). Altogether a catalogue of lesions underlying de-regulation of *MYC* and the NF-ΚB pathway, genomic instability in the form of APOBEC activity and chromothripsis, and t(4;14) emerged as risk factors for resistance to early intervention with triplet therapies and eventual disease evolution (**Supplemental Table 6**)^32^. Co-occurrence of many of these lesions, in parallel (i.e., genomic complexity), was a common theme amongst those with progressive disease. We finally assessed the effect on KRd/R outcome, considering any one or more of the [high-risk] features associated with treatment outcomes (**Supplemental Table 4**) and found that co-occurrence of any 2 features was associated with progression despite intervention with potent triplet therapy (p = 0.034; **Figure 4b**).

**Figure 3:**
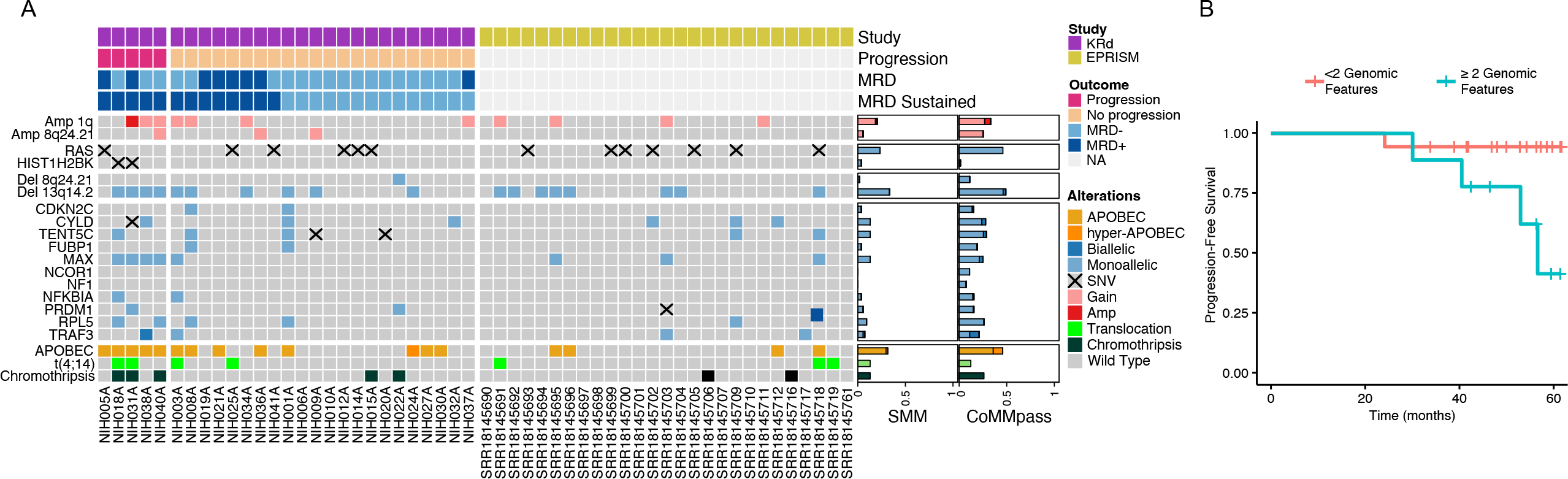
Genomic lesions associated with clinical outcomes in KRd/R-treated HR-SMM. **A**. Heatmap of genomic features found at significantly different frequency in comparison between NDMM and HR-SMM and within the KRd/R cohort associated with PFS or failure to sustain MRD-negativity. **B**. PFS curves stratified by presence of >1 features associated with PFS or failure to sustain MRD-negativity.

## DISCUSSION

With a growing armamentarium of active, tolerable therapies, there is increasing interest in the early treatment of patients with myeloma precursor disease. However, there is incongruity in currently available clinical risk stratification models for progression from SMM to MM^14^. In this study, to better understand the results of contemporary interventional studies of triplet therapies in the treatment of clinical HR-SMM, we used WGS and WXS to characterize the genomic landscape of HR-SMM in two parallel phase II interventional trials. We found that compared to NDMM, patients with HR-SMM had a lower frequency of established myeloma-defining genomic lesions; particularly those associated with poorer clinical outcomes and genomic complexity including a lower frequency and contribution of APOBEC-induced mutagenesis, and fewer disruptions of the *MYC* locus. Notably, these features are generally considered to be later events in the clonal evolution of disease progression to MM, but they are also two highly prevalent and powerful prognostic marker for predicting SMM progression^16-18, 32^. Though it could be argued that it would be expected for HR-SMM to be less genomically complex than MM, recent WGS studies have revealed that these late lesions are still generally acquired many years prior to clinically active disease and generally before the 2- to 5-year time-to-progression window expected for patients classified clinically as high-risk^6-9, 18, 19, 32^. Though the relative absence of these complex genomic features in the clinically defined HR-SMM cohorts does not imply that these patients would not have had early progression into MM in the absence of intervention, it does suggest that, at least at time of enrollment, they had genomically more indolent disease, or they were at lower risk of imminent progression in contrast to what is implied by their clinical risk scores, or a combination of both. In fact, patients on the KRd/R study who obtained deep and sustained responses (i.e. sustained MRD-negativity) and non-progressive disease appear genomically similar to patients with non-progressive monoclonal gammopathy and SMM under observation or to a small group of MM with very favorable outcomes (i.e., stable myeloma precursor conditions, **Supplemental Figure 5**)^17, 34^.

Among the overall genomic simplicity observed in treated clinically defined HR-SMM patients, there is a set of genomic features that portends sub-optimal outcome despite intervention. Combinations of APOBEC-induced mutagenesis, deregulation of *MYC* and the NF-ΚB pathway, and chromothripsis are associated with disease persistence (i.e., failure to achieve or sustain MRD-negativity), and progression from SMM to MM despite effective triplet therapy, as paralleled in treatment of NDMM. Whether these SMM patients with high genomic complexity would have had worse outcome without early intervention or whether early intervention changed the course of their disease are unanswered questions. Possibly, these lesions are representative of disease biology not susceptible to currently available interventional therapies provided on these clinical trials. For example, it has recently been seen that for patients with clinically defined high-risk SMM, even intensive early treatment with KRd followed by melphalan chemotherapy and autologous stem cell transplantation and maintenance therapy, only half are able to achieve MRD-negativity^35^. It seems reasonable to conjecture that myeloma precursor disease with genomically complex, high-risk disease biology (i.e., myeloma-to-be), may be better served by alternate diagnostic and novel therapeutic strategies. Future translational studies are needed to further address this unmet clinical need.

Taken together, given the elective nature of treatment of myeloma precursor conditions, our results support the use of genomic profiling to contextualize advantages of early intervention, to avoid overtreatment of non-progressors, and to better identify patients who are highly likely to progress in the absence of early intervention.

## ACKNOWLEDGEMENTS

This work was supported by the Myeloma Solutions Fund (MSF), Paula and Rodger Riney Multiple Myeloma Research Program Fund, the Tow Foundation, Sylvester Comprehensive Cancer Center NCI Core Grant (P30 CA 240139) and the Intramural NCI Program. B.D. is supported by the Sylvester K12 Calabresi Clinical Oncology Research Career Development Program. FM is supported by International Myeloma Society, NIH, and American Society of Hematology.

## AUTHOR CONTRIBUTIONS

D.K. and O.L. designed and supervised the clinical trial, collected the data and wrote the paper. B.D and F.M. designed and supervised the genomic correlative study, collected, and analyzed the data and wrote the paper. M.P. collected and analyzed the data and wrote the paper. P.B., M.C., and M.D. collected and analyzed the data. E.H. and R.S.P designed and collected the data. K.M., R.Y., S.U., G.G., I.G., and G.M collected the data.

All authors read, revised, and proofed the manuscript.

## CONFLICT OF INTEREST

B.D. has received honoraria from advisory boards from Sanofi and Janssen and Independent Data Review Committee for Janssen.

D.K. acknowledges research funding from: NCI/NIH, FDA, MMRF, DoD-PROMETHEUS (Murtha Cancer Center Research Program), Amgen, Celgene, Janssen, Karyopharm; has received honoraria for advisory boards and presentations for: Alphasights, Aptitude Health, Arcellx, BMS, Bridger Consulting Group, Curio Science, Karyopharm, MJH Life Sciences, MMRF, Plexus Communications, Sanofi; and served on Independent Data Monitoring Committees (IDMC) for: Aperture Medical Technology, Arcellx.

O.L. acknowledges research funding from: NCI/NIH, FDA, LLS, Rising Tide Foundation, Memorial Sloan Kettering Cancer Center, MMRF, IMF, Paula and Rodger Riney Foundation, Myeloma Solutions Fund, Perelman Family Foundation, Amgen, Celgene, Janssen, Takeda, Glenmark, Seattle Genetics, Karyopharm; has received honoraria for scientific talks/participated in advisory boards for: Adaptive, Amgen, Binding Site, BMS, Celgene, Cellectis, Glenmark, Janssen, Juno, Pfizer; and served on Independent Data Monitoring Committees (IDMC) for international randomized trials by: Takeda, Merck, Janssen, Theradex.

S.U. acknowledges research funding from Amgen, Array Biopharma, BMS, Celgene, GSK, Janssen, Merck, Pharmacyclics, Sanofi, Seattle Genetics, SkylineDX, Takeda; honoraria/consulting from Abbvie, Amgen, BMS, Celgene, EdoPharma, Genentech, Gilead, GSK, Janssen, Oncopeptides, Sanofi, Seattle Genetics, SecuraBio, SkylineDX, Takeda, TeneoBio; grant support from NCI, LLS, MMRF.

None of the other Authors have conflict of interest to disclose.

## DATA AVAILABILITY

The dataset used for this paper is derived from public and newly sequenced sources: 27 SMM exomes were imported from dbGaP: phs001323.v3.p1. 701 MM exomes (from patients with RNA-seq and low coverage WGS data) were imported from the CoMMpass trial; IA 13. Data from the 27 newly sequenced samples are currently uploading on EGA and will be available upon publication of the manuscript. KRd +/- Dara: EGAS00001007404.

## CODE AVAILABILITY

Analyses conducted using previously published code are detailed and available via github links in **Supplemental Methods**.

## Notes

### Author Declarations

Samples and data were obtained and managed in accordance with the Declaration of Helsinki and the Institutional Review Board of the University of Miami under protocol 20210398.

