## Supplemental Information for "Genomic Profiling to Contextualize the Results of Intervention for High-Risk Smoldering Myeloma"

#### SUPPLEMENTAL METHODS

##### Sample Preparation

For patients on NCT01572480, available samples from pre-treatment bone marrow were thawed and CD138+ microbeads were used to sort plasma cells from frozen bone marrow mononuclear cells. Normal match was selected for each patient from peripheral blood mononuclear cells.

##### Sequencing and Analytical Methods

Sequencing was performed at New York Genome Center. Following quantification via PicoGreen and quality control by Agilent Bioanalyzer, ~500 ng of genomic DNA was sheared (LE220-plus Focused-ultrasonicator; Covaris, catalog no., 500569) and sequencing libraries were prepared using a modified KAPA Hyper Prep Kit (Kapa Biosystems, KK8504). Briefly, libraries were subjected to a 0.5 × size select using aMPure XP Beads (Beckman Coulter, catalog no., A63882) after post-ligation cleanup. Libraries that were not amplified by PCR (07652_C) were pooled equivolume. Libraries amplified with five cycles of PCR (07652_D, 07652_F, and 07652_G) were pooled equimolar. Samples were run on a NovaSeq 6000 in a 150 bp/150 bp paired-end run, using the NovaSeq 6000 SBS v1 kit and an S4 Flow Cell (Illumina), as described previously[1]. Target coverage depth was 80x for tumor and 40x for normal.

**Whole-genome analysis pipeline.** Coverage for tumor and normal samples are reported in **Extended Data Table 1**. Short insert paired-end reads were aligned to the reference genome (GRCh38) using the Burrows–Wheeler Aligner (v0.5.9; ref. 17). All samples were uniformly analyzed by the following bioinformatic tools: somatic mutations were identified by MuTect2, VarScan2, and Strelka[2-4]; indels were identified by VarScan2, MuTect2, SvABA, and Strelka[2-5]; copy number analysis and tumor purity (i.e., cancer cell fraction) were evaluated using ASCAT[6]; IgCaller was used to identify translocations at the immunoglobulin loci[7]; structural variants were defined by Manta, DELLY, and SvABA[5, 8, 9] and passed through additional quality filters, and were manually curated to define complex events (i.e., templated insertions, chromothripsis, and chromoplexy) as described previously[10]. Chromothripsis was called from WXS data, additionally, using methods described by Maclachlan et al[11]. SV hotspots were identified and called using previously described methods[12]. An SV hotspot was considered to be involved by structural variation in a given sample if either an SV breakpoint fell within the hotspot or withing 1MB of its start or end. We also considered effect and impact. For example: a deleterious SV within an amplification hotspot would not be considered.

The exomes data downloaded from public repositories were aligned to the reference human genome (GRCh37) using Burrows-Wheeler Aligner, BWA (v0.7.17). Deduplicated aligned BAM files were analyzed using FACETS (v0.5.6, https://github.com/mskcc/facets) for copy number variants, and otherwise subjected to the same pipeline as for genomes above – with the exception of structural variant calling.

**Mutational Signatures.** Mutational signatures were analyzed across all whole genomes. To estimate the activity of mutational signatures, we first employed a three step process of de novo extraction, assignment, and fitting[13]. For the first step, we ran SigProfiler for SBS signatures[14]. All extracted signatures were then compared with the latest Catalogue of Somatic Mutations in Cancer (COSMIC) reference (https://cancer.sanger.ac.uk/cosmic/signatures/SBS) to identify the known mutational processes active in the cohort. For SBS, we applied mmsig (https://github.com/UM-Myeloma-Genomics/mmsig)[15], a fitting algorithm, to confirm the presence and estimate the contribution of each mutational signature in each sample guided by the catalog of signatures extracted for each individual sample by SigProfiler's de novo refit.

For unbiased comparisons of mutational signatures, HR-SMM WGS were compared to WGS from patients with NDMM treated with KRd +/- Dara[16]. SBS2 and SBS13 contribution were summed across WGS samples and hyper-APOBEC samples were defined as those with expression above the 10$^{th}$ decile. The same analysis for HR-SMM WXS and CoMMpass WXS data was performed.

**Copy Number Aberrations and Driver Mutations**

Given the size of the SMM cohort, GISTIC peak discovery was not performed. Instead, reference GISTIC peaks previously identified as significant in MM were used as reference[12]. The analysis was executed using Gene Pattern web interface (http://genepattern.broadinstitute.org)

We used the dN/dScv package to annotate genes in our cohort[17]. Given the size of the SMM cohort, driver discovery was not performed. Instead, a reference driver gene list was used to designate driver status to identified variants[18]. Similarly, SV hotspots, recurrent CNV and GISTIC peaks were referenced from prior work[12, 18].

We then developed a locus-based classification scheme, combining all of focal and large CNA, GISTIC peaks, and SNV to detect dysregulation at driver loci[18]. These methods allow for identification of common biological processes affected by various genomic aberrations (https://github.com/UM-Myeloma-Genomics/GCP_MM). CNA were used as a proxy for SV given the combination of both WGS and WXS in this study. Sensitivity analysis was performed with removal of 2 HR-SMM cases that would be reclassified as MM per IMWG 2014 diagnostic criteria[19]. Overall and progression-free survival annotations were pulled from CoMMpass clinical annotations[18].

To transcend constraints of clinically applied disease definitions, clinical annotations were removed, and cases were clustered together (701 NDMM and 54 HR-SMM) to determine the relationship of genomic lesions to underlying disease state. We used the R Package, pheatmap, to perform hierarchical clustering to report on patterns of co-occurrence (https://github.com/raivokolde/pheatmap).

1. Jones, D., K.M. Raine, H. Davies, P.S. Tarpey, A.P. Butler, J.W. Teague, et al., *cgpCaVEManWrapper: simple execution of CaVEMan in order to detect somatic single*

*nucleotide variants in NGS data.* Current protocols in bioinformatics, 2016. **56**(1): p. 15.10. 1-15.10. 18.

### Genomic Profiling to Contextualize the Results of Intervention for High-Risk Smoldering Myeloma

#### Extended Data Figures

#### Extended Data Figure 1. Mutational Signatures Landscape of WGS samples taken from NDMM patients treated with either of KRd or Dara-KRd.

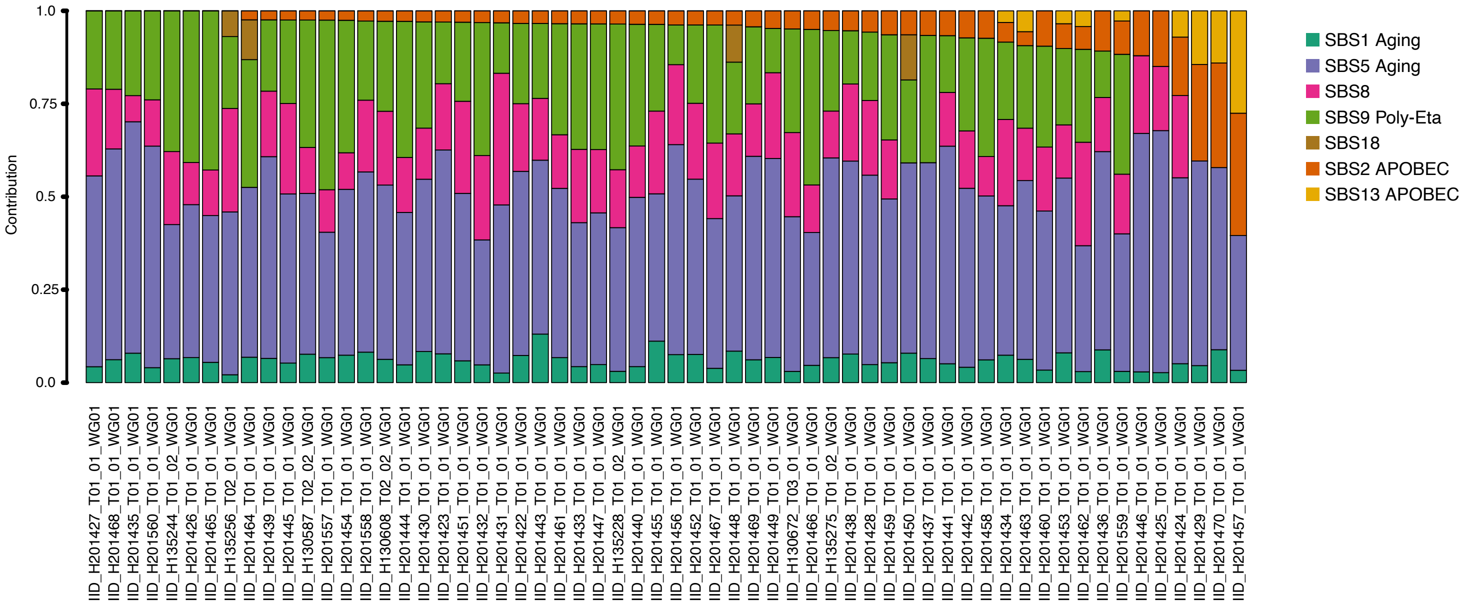

**Extended Data Figure 2: Cumulative Copy Number Profiles for the treated HR-SMM cohort (n= 54; A) and NDMM from CoMMpass (n=701; B).**

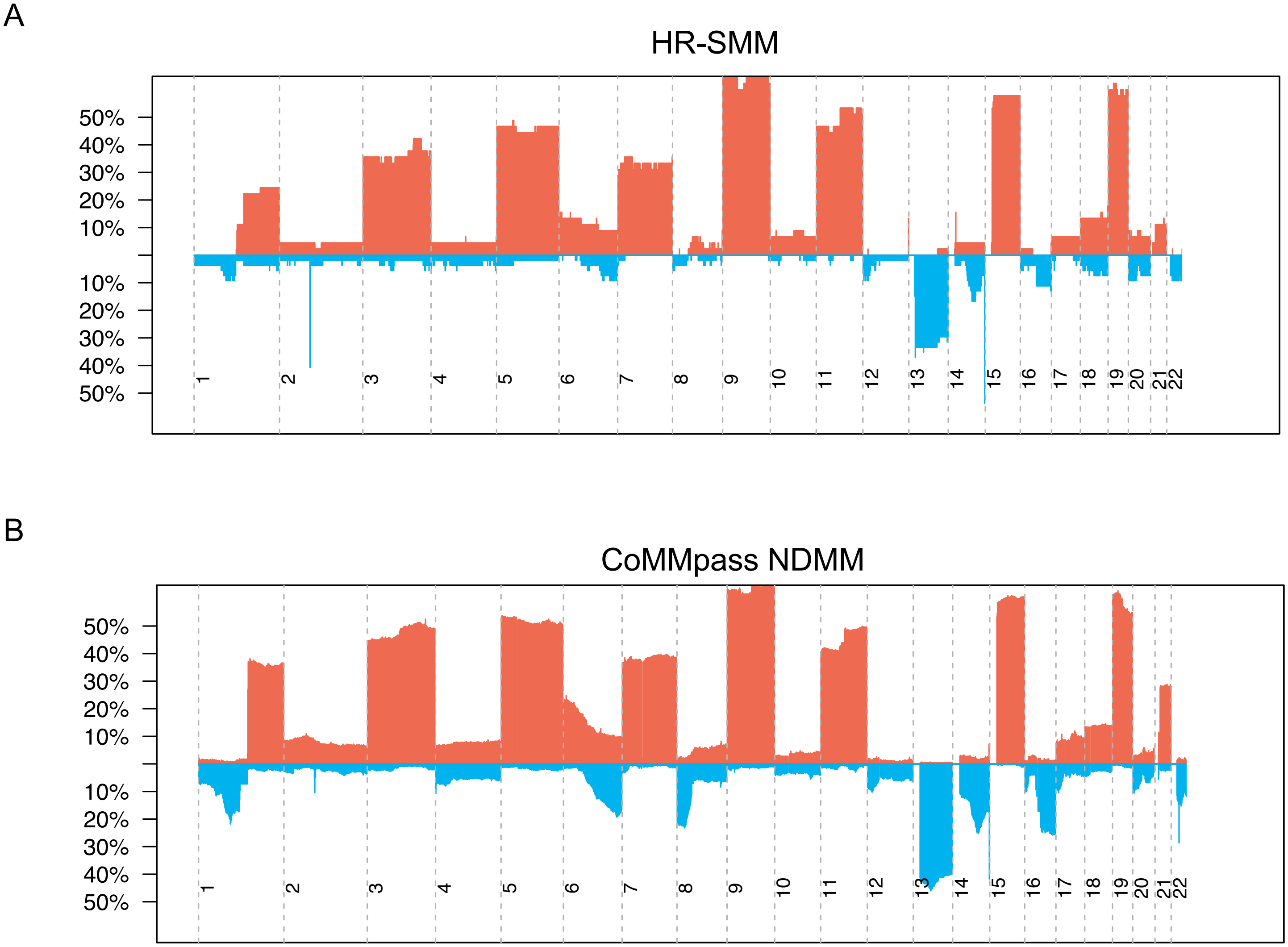

**Extended Data Figure 3: Heatmap of genomic features assessed in comparison between HR-SMM and NDMM.** Features are those assessable combining mutational and copy number data to facilitate comparison of alternate sequencing technologies.

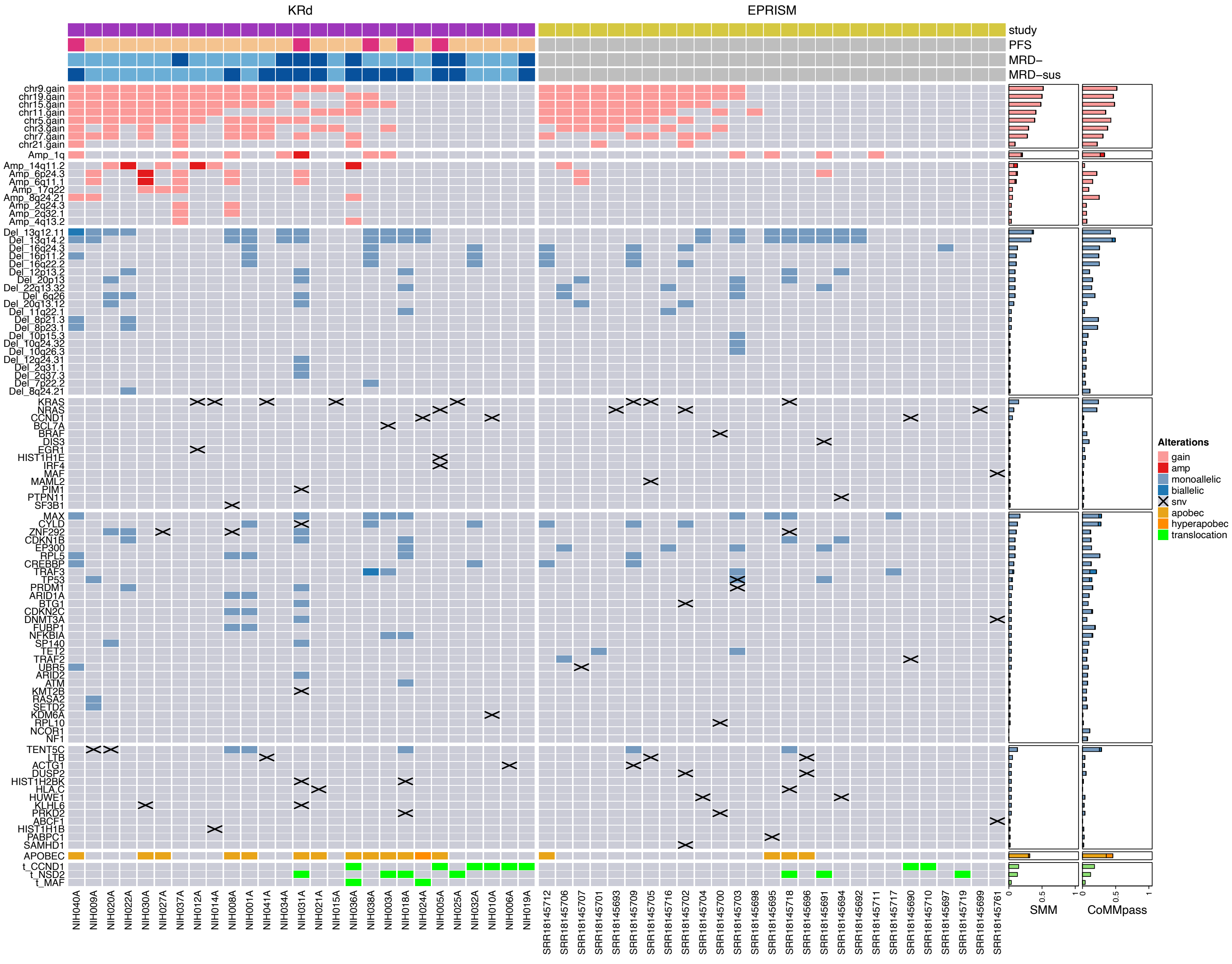

**Extended Data Figure 4: Comparison of Structural Variant Hotspots between KRd/R-treated HR-SMM and KRd +/- Dara-treated NDMM whole genomes (A). B. Total SV hotspots between KRd/R-treated HR-SMM and KRd +/- Dara-treated NDMM whole genomes. C. Total SV hotspots between HR-SMM Progressors and Non-Progressors after treatment with KRd/R.**

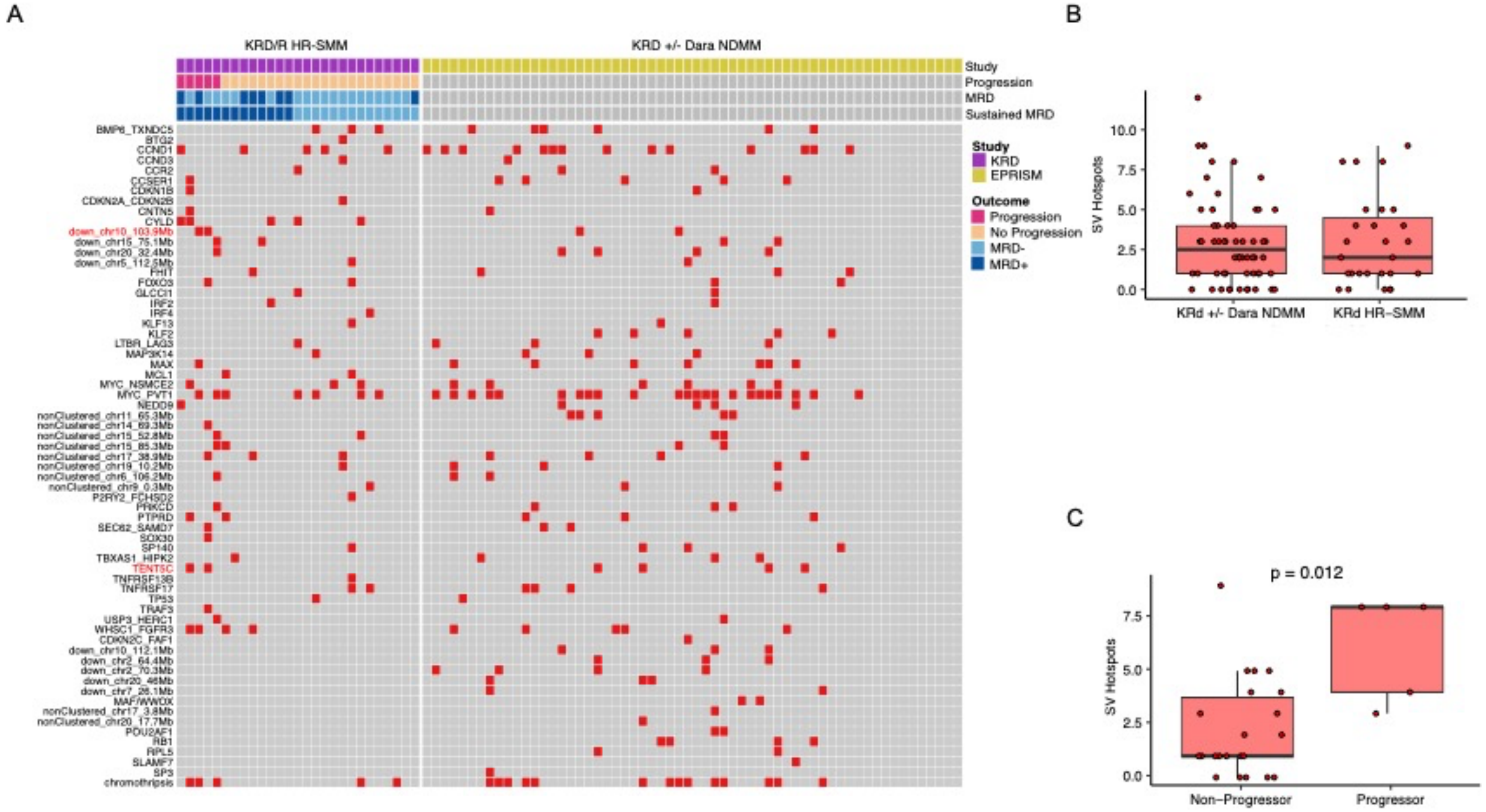

**Extended Data Figure 5: Hierarchical clustering based on genomic features of all HR-SMM (n=54) and NDMM from CoMMpass (n=701).**

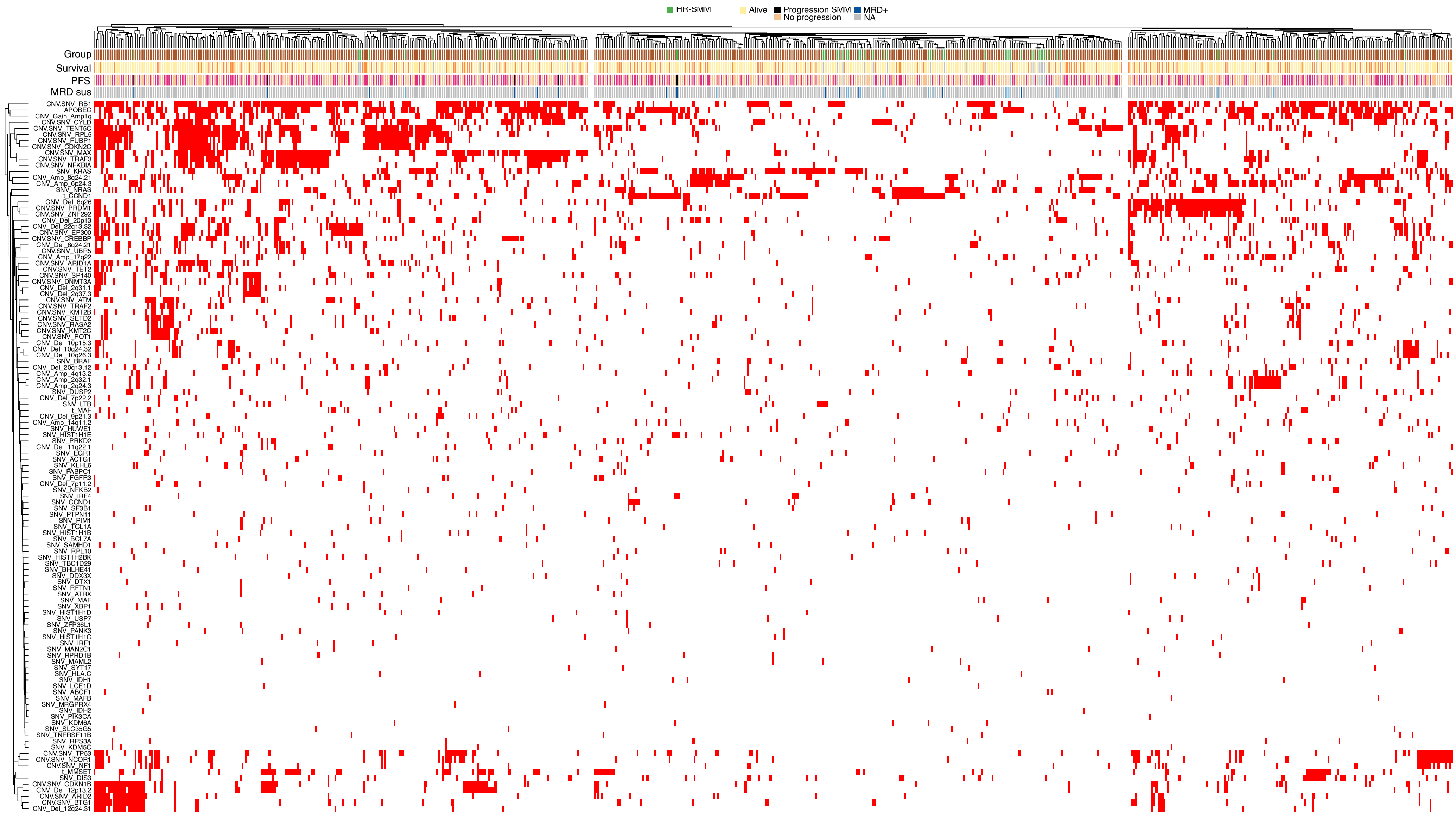

#### Extended Data Figure 6: Circos Plots for KRd/R Patients. Colored sample text represents patients with progression.

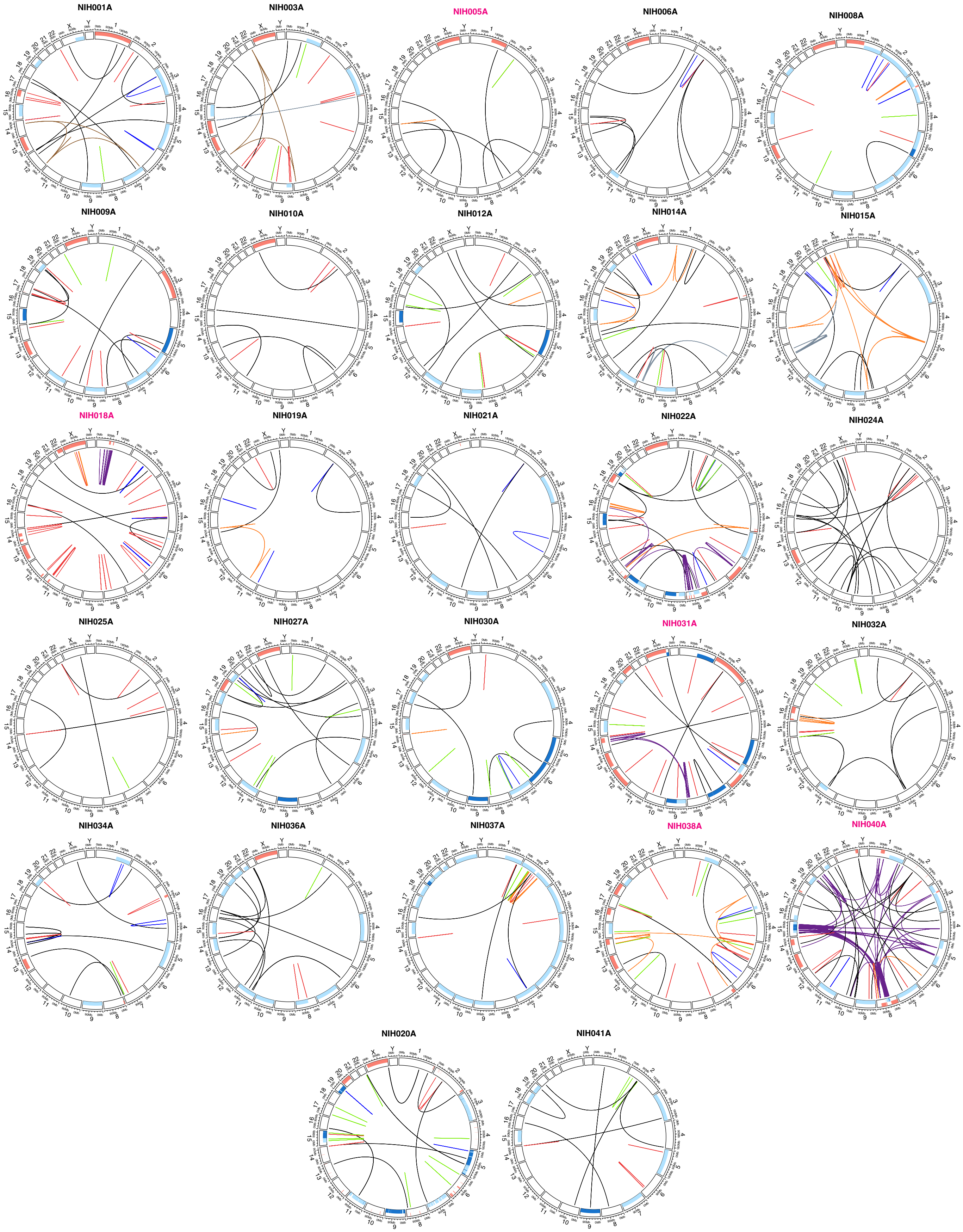
